# LCatlas: A Point-Of-Care Tool for Predicting Immunotherapy Outcomes in Non-Small Cell Lung Cancer

**DOI:** 10.64898/2026.11.18.26368446

**Authors:** Sung Yong Park, Alexis Leyba, Wilson Yeung, Robert C Hsu, Jorge J Nieva, Ha Youn Lee

## Abstract

Non-small cell lung cancer (NSCLC) remains a leading cause of cancer mortality, with a minority of patients experiencing durable responses to first-line immune checkpoint inhibitors (ICIs). Currently, there are no easily accessible tools to predict clinical benefit to first-line immunotherapy. To address this gap, we developed LCatlas (https://www.lcatlas.org), a point-of-care web application designed to predict immunotherapy outcomes and inform evidence-based treatment decisions. Using data from 79 NSCLC patients receiving first-line immunotherapy, we extracted demographic, clinical, and molecular features from electronic medical records and compared three independent approaches–statistical model (logistic regression), machine learning (XGboost), and AI model (TabTransformer) to identify the most effective predictor. LCatlas accepts nine variables—gender, smoking history, bone/pleura/liver metastasis, PD-L1 tumor staining, albumin level, hemoglobin level, lymphocyte count, neutrophil count, and platelet count–to predict responses to both immunotherapy and immunotherapy plus chemotherapy. LCatlas cross-validation yielded an area under the curve (AUC) of 79% [72% - 85%], with sensitivity of 87% [72% - 95%] and specificity of 78% [64% - 89%], outperforming previous studies including high-volume data-driven deep-learning approaches. In a validation cohort consisting of 28 individuals, it demonstrated a prediction accuracy of 82%. LCatlas may serve as a valuable point-of-care tool by identifying patients likely to experience extended immunotherapy or combination therapy using readily available medical data. Its evidence-based approach can optimize personalized treatment planning and thereby promote quality and safety of care.

## INTRODUCTION

In 2023, the global incidence of non-small cell lung cancer (NSCLC) surpassed 2 million new diagnoses [1]. The disease remains the leading cause of cancer-related mortality, with a 5-year overall survival (OS) of approximately 25% [2, 3]. Over the past decade, immune checkpoint inhibitors (ICIs) targeting the PD-1/PD-L1 axis have transformed the therapeutic landscape of NSCLC. First-line monotherapy with pembrolizumab or nivolumab has achieved durable responses in NSCLC patients across all stages [4, 5], while combination regimens of ICIs with platinum-based chemotherapy or anti-CTLA-4 agents have expanded the indications to a broader patient population [6, 7]. Survival in NSCLC is impacted by factors such as stage at diagnosis, histology type, the presence of actionable driver mutations and PD-L1 status. In patients without an actionable driver mutation, progression-free survival remains between 6 – 10 months for individuals with advanced NSCLC treated with first line therapy [8]. However, these drugs carry inherent toxicity, and shared decision making with patients requires accurate probabilistic estimation of long-term benefit against these risks. Consequently, identifying patients likely to respond long term to immunotherapy is a critical clinical need, and clinical decision support systems [9] that identify long term responders would be valuable.

Current biomarkers – PD-L1 expression, tumor mutational burden (TMB), and circulating tumor DNA (ctDNA) dynamics – are imperfect, often conflicting, and not routinely available in all clinical settings [10–12]. The heterogeneity of NSCLC at the genomic, transcriptomic, and immunologic levels poses a major challenge to the identification of reliable predictors of immunotherapy response [13, 14]. Various deep learning and machine learning models have been used to uncover latent patterns associated with treatment outcomes from high-dimensional, multimodal data encompassing CT images, blood cytokine profiles, genomic information, and clinical data [15–19]. However, there are no readily accessible point-of-care tools to help physicians predict immunotherapy response using routinely collected data in clinical practice. Additionally, no clinical decision support system currently aids in choosing between first-line immunotherapy alone and immunotherapy plus chemotherapy. Currently, no prospective head-to-head trial definitively favors one strategy over the other and treatment selection remains guided by clinical context rather than biomarker threshold alone [20, 21].

To address gaps in predicting response to first-line therapy, we developed LCatlas (Lung Cancer atlas), a user-friendly web tool that predicts responses to first-line immunotherapy and combination therapy (immunotherapy and chemotherapy). LCatlas uses a curated set of nine routinely collected demographic, clinical, and molecular variables. We assessed the accuracy of three independent approaches – logistic regression, a machine learning algorithm, and an AI model – applied to data from 79 NSCLC patients who initiated treatment between 2016 and 2023, selecting the prediction algorithm for LCatlas. Its predictive performance was validated in a separate cohort of 28 patients treated between 2020 and 2025. Our goal is to provide point-of-care software that identifies patients likely to benefit from immunotherapy or combination therapy, leading to prompt, personalized treatment and improved outcomes by leveraging readily available medical records.

## MATERIALS AND METHODS

### Study population and data collection

This retrospective study included patients with pathologically confirmed NSCLC treated at the Los Angeles General Medical Center and the University of Southern California Norris Comprehensive Cancer Center [22]. This study was approved by the University of Southern California Institutional Review Board (IRB HS-15-00200). Given the retrospective nature of the study, informed consent was waived. Data were obtained from 79 patients’ electronic medical records via retrospective chart review between January 2015 to June 2023. These records were accessed for research purposes from January 10, 2025 to May 19, 2026 and no identifiable information were accessed. These patients received first-line immunotherapy, either alone or in combination with other therapies. Twenty-nine patients (37%) received immunotherapy only, while 45 (57%) received immunotherapy combined with chemotherapy. Five patients received investigational agents in combination with immunotherapy.

Collected data included demographic information and clinical characteristics at diagnosis or initiation of immunotherapy (age, gender, race/ethnicity, smoking history, Body Mass Index (BMI), clinical stage, Eastern Cooperative Oncology Group (ECOG) performance score, histology, metastasis, and first-line therapy type). Race/ethnicity was self-reported; smoking history was extracted from chart review. Results of molecular testing (PD-L1 tumor staining percentage, albumin level, hemoglobin level, lymphocyte count, neutrophil count, and platelet count) and genomic profiling, were obtained from testing reports (see [22] for more details).

At treatment initiation, 85% of patients had stage IV disease. Ten patients had stage III and one each at stages I and II. ECOG performance statuses ranged from 0 to 2. The age distribution was as follows: 6 patients were over 80 years old, 19 were between 71 and 80, 29 between 61 and 70, 18 between 51 and 60, 5 between 41 and 50, and 2 patients were between 21 and 30 years old. The cohort consisted of 51 males (65%) and 28 females (35%). Ethnic groups represented included 33 Asian, 14 Hispanic, 17 White, 6 Black, 6 Armenian, 2 European, and 1 patient of other ethnicity. The percentage of missing data for each variable is listed in Table 1, denoted as ’NA’. Patients with *ALK*, *EGFR*, and *ROS1* mutations were excluded, as first-line immunotherapy is generally not recommended for NSCLC patients with these mutations [23].

**Table 1.** Predictors used in LCatlas variable selection and odds ratios with 95% confidence intervals. *condition1: immunotherapy alone with PD-L1 ≥50% or combination therapy with PD-L1 <50% *condition2: immunotherapy alone with PDL1 <50% or combination therapy with PD-L1 ≥50%

| **Predictors** | **AUC** | **Sensitivity, Specificity** | **Cohort distributions** | **Odds ratio** |
|---|---|---|---|---|
| Treatment type + PD-L1 | 55% | 87%, 24% | condition1<sup>*</sup> (68.4%), condition2<sup>*</sup> (19.0%), NA (12.7%) | 0.39 [0.04 – 3.50] |
| Albumin | 78% | 80%, 72% | High (34.2%), Low (51.9%), NA (13.9%) | 16.11 [2.78 – 93.57] |
| SII (Systemic Immune-inflammation Index) | 72% | 54%, 89% | High (70.9%), Low(16.5%), NA (12.7%) | 0.25 [0.038 – 1.62] |
| Bone/Pleura/Liver metastasis | 71% | 58%, 83% | No (20.3%), Yes (60.8%), NA (19.0%) | 0.27 [0.05 – 1.55] |
| Hemoglobin | 62% | 88%, 40% | High (63.3%), Low (31.6%), NA (5.1%) | 3.07 [0.29 – 33.03] |
| Gender | 61% | 82%, 40% | Male (64.6%), Female (35.4%), NA (0%) | 0.16 [0.03 – 1.02] |
| Smoking history | 59% | 94%, 24% | Never (20.3%), Former or current (79.7%), NA (0%) | 2.93 [0.24 – 36.26] |
| BMI | 59% | 63%, 58% | Low (50.6%), High (44.3%), NA (5.1%) | Not included |
| *KRAS* mutation | 56% | 88%, 24% | No (74.7%), Yes (20.3%), NA (5.1%) | Not included |
| Stage | 52% | 18%, 86% | III or lower (15.2%), IV (84.8%), NA (0%) | Not included |
| ECOG | 51% | 88%, 13% | 0 or 1 (87.3%), 2 (12.7%), NA (0%) | Not included |

From a separate validation cohort of 28 NSCLC patients, the LCatlas input variables – gender, smoking history, bone/pleura/liver metastasis, PD-L1 tumor staining percentage, albumin level, hemoglobin level, lymphocyte count, neutrophil count, and platelet count — were collected from their medical records (Supplementary Table S1). These individuals initiated first-line immunotherapy, either alone or in combination with chemotherapy, between December 2020 and March 2025.

### Logistic regression

We used logistic regression (R, version 4.1.2, RRID:SCR_001905) to predict immunotherapy outcome in NSCLC patients based on demographic, clinical, and molecular variables. The primary outcome was a binary classification of time to failure, categorized as less than or greater than one year. Lymphocyte count, neutrophil count, and platelet count were combined into a single variable of the systemic immune-inflammation index (SII), calculated as neutrophil-to-lymphocyte ratio (NLR) multiplied by platelet count [24, 25]. Bone, Pleura, Liver metastasis statuses were combined into a single variable (Yes: presence of metastasis in one or more of these organs; No: none).

The first predictor was a composite of first-line therapy type (immunotherapy or immunotherapy plus chemotherapy) and PD-L1 tumor staging percentage, defined as: condition1: immunotherapy alone with PD-L1 tumor staining ≥50% or immunotherapy and chemotherapy with PD-L1 tumor staining <50%; condition2: immunotherapy alone with PD-L1 tumor staining <50% or immunotherapy and chemotherapy with PD-L1 tumor staining ≥50%. Albumin level, SII, bone/pleura/liver metastasis, hemoglobin level, gender, smoking history, BMI, KRAS mutation status, clinical stage and ECOG score were categorized, as presented in Table 1. Single variable prediction accuracy was assessed using the AUC for each of the 11 categorical variables (Table 1). Sensitivity was the proportion of patients with extended benefit (greater than one year) correctly classified, and specificity was the proportion of those with early progression (less than one year) correctly identified. Optimal thresholds for albumin, SII, hemoglobin, and BMI were determined by Receiver Operating Characteristic (ROC) curve analysis, using Youden’s J statistic to maximize the combined sensitivity and specificity.

Missing values were imputed using the Multivariate Imputation by Chained Equations (MICE) method using the ‘mice’ R package with predictive mean matching (PMM) (RRID:SCR_001905). The outcome variable was a binary indicator of time on treatment exceeding a one-year threshold. A logistic regression model was fitted to data from 79 NSCLC patients’ data using the ‘*glm*’ function with a logit link.

Model performance was evaluated using AUC, sensitivity, specificity, and accuracy. We used cross-validation to identify the optimal model. Beginning with the composite variable of treatment type and PD-L1 tumor staining percentage, we incrementally added predictors ranked by AUC values and evaluated cross-validation accuracy with each addition. LCatlas predictors were identified by maximizing AUC, averaged across cross-validation tests; average performance metrics along with 95% confidence intervals were determined by performing 100 runs of 4-fold cross-validation. To prevent data leakage, MICE was performed within each cross-validation fold; the imputation model was fitted using only the training data and subsequently applied to the test data. The Akaike Information Criterion (AIC) was also evaluated as the number of predictors changed.

### Xgboost

We used XGBoost (version 0.90, RRID:SCR_021361), a machine learning algorithm, to predict time to immunotherapy failure [26]. The input consisted of the categorial variables defined above, and the outcome was treatment failure within one year or greater than one year. Missing values were imputed as described above. An XGBoost classifier was trained using 1,000 estimators and a learning rate of 0.01. Model performance was evaluated by 100 iterations of 4-fold cross-validation, reporting mean AUC, sensitivity, specificity, accuracy and 95% confidence intervals.

### AI model

We used an AI model built with TensorFlow and Keras (RRID:SCR_026159)—a TabTransformer—to predict time-to-failure using the 11 categorial variables defined above. Missing values were imputed as previously explained. The architecture included one Transformer block, composed of Layer-Normalization, Multi-Head Attention, Dropout, a feed-forward sub-network. The output from the Transformer block was flattened and then passed through a Multi-Layer Perceptron (MLP) consisting of three fully connected layers with 64, 32, and 16 units, respectively. Dropout with a rate of 0.2 was applied after the first two dense layers to reduce overfitting. The final output layer consisted of a single neuron with a sigmoid activation function to produce a probability estimate for binary classification of treatment failure within one year or greater.

## RESULTS

### Clinical and demographic variables and immunotherapy outcomes

Among 79 patients who received first-line immunotherapy, either alone or in combination with chemotherapy or others, time on treatment exceeded one year for 17 patients (21.5%) and was less than one year for 62 patients (78.5%) (Figure 1A). Clinical stage at immunotherapy initiation was not significantly associated with time to treatment failure (Figure 1B), and its AUC was low at 52% with 18% sensitivity and 86% specificity (Table 1). Similarly, immunotherapy responses did not differ significantly between NSCLC subtypes, adenocarcinomas and others (Figure 1C). We found no significant associations between treatment outcome and age (Figure 1D).

**Figure 1.**
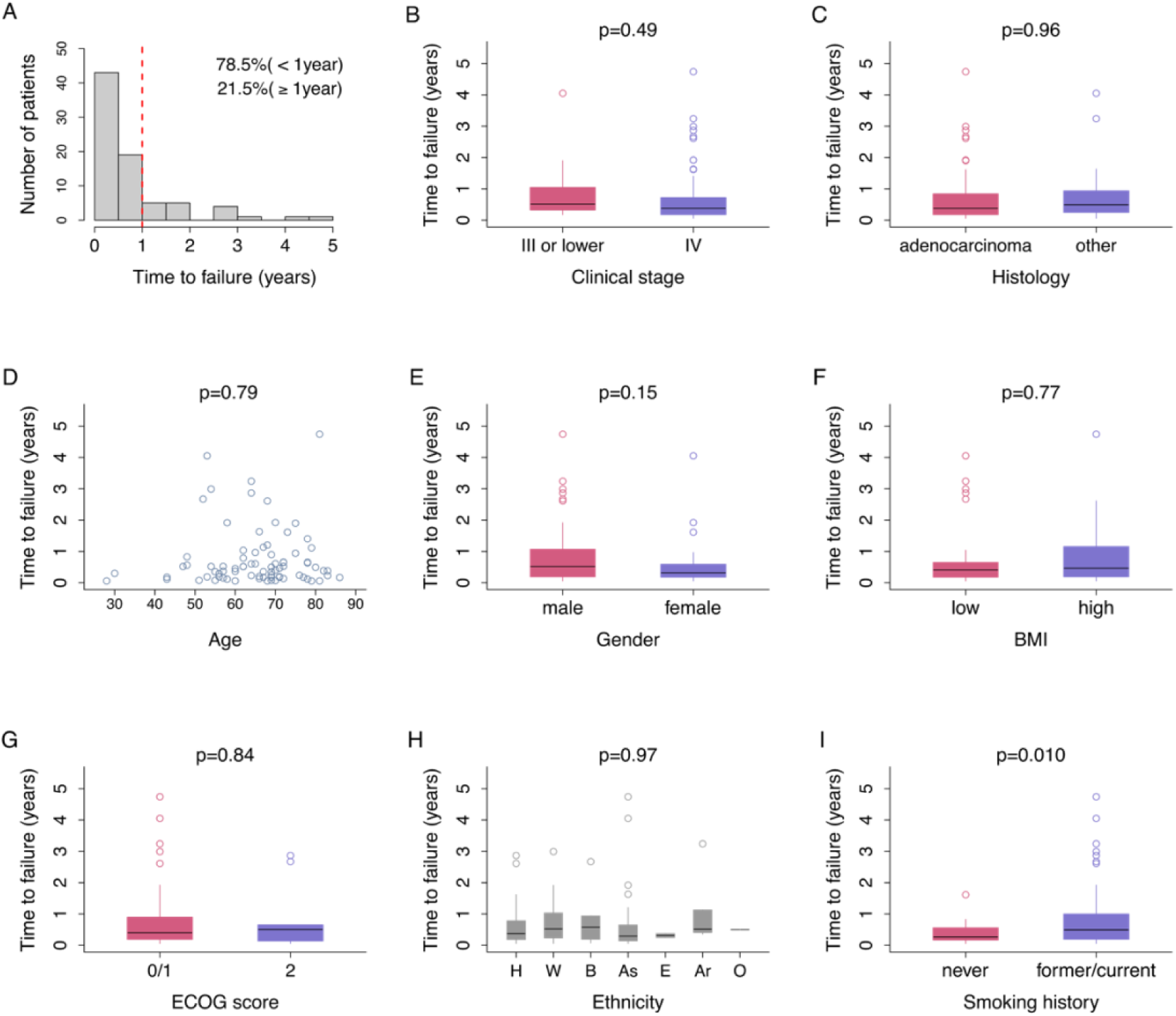
Time to treatment failure and patient characteristics. **A.** Distribution of time to treatment failure. Among 79 NSCLC patients. 21.5% (n=17) remained on treatment for over one year. **B.** Comparison of time to treatment failure stratified by disease stage. Patients with stage III or lower NSCLC (n=12) had a mean time to treatment failure of 0.96 [95% CI, 0.27 – 1.65] years, whereas those with stage IV NSCLC (n=67) had a mean time to treatment failure of 0.72 [0.50 – 0.95] years (t-test: p=0.49). **C.** Boxplot comparing time to treatment failure for patients with adenocarcinoma NSCLC (n=48) and those with others including squamous cell carcinoma NSCLC (n=29), with mean times to treatment failure for 0.77 [0.48 – 1.06] years and 0.77 [0.43 – 1.12] years, respectively. **D.** Scatter plot of age and time to failure in 79 NSCLC patients. The Pearson correlation coefficient was 0.031 (t-test; p=0.79). **E.** Boxplot comparing time to treatment failure for male (n=51) and female (n=28) patients. Time to failure was longer for male patients (0.87 [0.59 – 1.15] years) compared to female patients (0.56 [0.25 – 0.88] years). **F.** Time to immunotherapy failure in patients with high (≥ 25) BMI (n=35) versus those with low (< 25) BMI (n=40). The mean time to failure was 0.81 [0.48 – 1.13] years in the high BMI group and 0.74 [0.43 – 1.05] years in the group with low BMI. **G.** Comparison of time to failure between patients with ECOG scores of 0 or 1 (n=69) and those with an ECOG score of 2 (n=10). Time to failure difference was not significant (0.75 [0.53 – 0.98] vs. 0.82 [0.07 – 1.58] years; t-test, p=0.84). **H.** Comparison of time to failure across different ethnic groups: Hispanic (n=14), White (n=17), Black (n=6), Asian (n=33), European (n=2), Armenian (n=6), and Other (n=1). **I.** Comparison of time to immunotherapy failure among NSCLC patients with different smoking histories: never smokers (n=16) and former or current smokers (n=63). Never smokers had significantly shorter times to immunotherapy failure (0.42 [0.21 – 0.63] years) compared to former or current smokers (0.85 [0.59 – 1.10] years).

On average, male patients had a longer time to failure than female patients (Figure 1E). A higher proportion of patients with a BMI of 25 remained on treatment for over one year compared to those with lower BMIs (29% vs. 15%); however, this difference was not statistically different (Figure 1F).

Patients with ECOG scores of 0 or 1 had similar times to failure with those with ECOG scores of 2 (Figure 1G). The duration of immunotherapy for each ethnic group is presented in Figure 1H. Patients with a history of smoking – including both former and current smokers – had significantly longer time on treatment compared to those with no smoking history (Figure 1I).

### Impact of PD-L1, therapy type and blood biomarkers on treatment outcomes

Whether patients received first-line immunotherapy alone or in combination with chemotherapy did not influence treatment outcomes (Figure 2A). Tumor proportion score (TPS) measured by PD-L1 tumor staining percentage was not associated with immunotherapy outcome (Figure 2B). We observed similar distributions of time to failure across the three PD-L1 tumor staining percentage groups (0%, 1-49%, and 50-100%; Supplementary Figure S1). However, time to immunotherapy failure was affected by a combination of first-line therapy type and PD-L1 tumor staining percentage. As shown in Figures 2C and 2D, patients receiving first-line immunotherapy alone with PD-L≥50%, or immunotherapy plus chemotherapy with PD-L1<50% had longer times to failure than those receiving the reverse combinations (immunotherapy alone with PD-L1<50% or immunotherapy plus chemotherapy with PD-L1≥50%). This composite variable of treatment type plus PD-L1 tumor staining percentage showed an AUC of 55% with 87% sensitivity and 24% specificity. However, the number of patients in the reverse combination group (n=15) was small compared to the first group (n=54).

**Figure 2.**
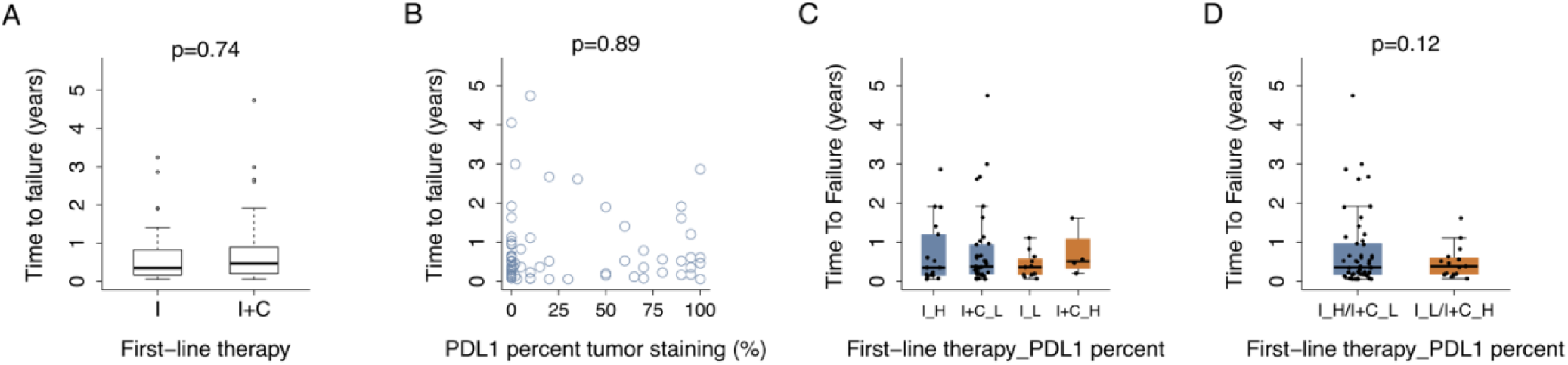
Treatment response, first-line therapy type and PD-L1 tumor staining. **A.** Comparison of time to immunotherapy failure between patients who received immunotherapy only (I) as their first line therapy (n=29, 0.69 [0.37 – 1.01] years) and patients who received immunotherapy plus chemotherapy (I+C) as their initial therapy (n=45, 0.76 [0.48 – 1.04] years; t-test, p=0.74). **B**. Scatter plot of time to immunotherapy failure and percentage of PD-L1 staining in tumor cells, at the start of treatment among 74 patients. The Pearson correlation coefficient was 0.016 (p=0.89). **C**. Time to failure across four groups defined by treatment and PD-L1 tumor proportion score: (1) Immunotherapy alone with PD-L1 tumor staining ≥50% (I_H, n=17), (2) Immunotherapy plus chemotherapy with PDL1 <50% (I+C_L, n=37), (3) Immunotherapy alone with PD-L1 <50% (I_L, n=11), and (4) Immunotherapy plus chemotherapy with PD-L1 ≥50% (I+C_H, n=4). **D.** Comparison between the first two groups (I_H and I+C_L) and the latter two groups (I_L and I+C_H). The mean time to failure was 0.76 [0.50 – 1.02] and 0.49 [0.26 – 0.73] years, respectively.

Lymphocyte counts at the onset of immunotherapy were significantly associated with longer time to treatment failure (Figure 3A). Conversely, the correlation coefficient between neutrophil count and time to failure was negative (Figure 3B). Consequently, the correlation coefficient between neutrophil-lymphocyte ratio (NLR) and time to failure was negative (Supplementary Figure S2). A negative correlation coefficient was observed between platelet levels and time to failure (Figure 3C). Combining these variables, the systemic immune-inflammation index (SII), defined as NLR multiplied by platelet count, demonstrated high single variable accuracy with an AUC of 72%. Patients with SII lower than 504 ×10^9^ cells/L had significantly greater times to failure than those with higher SII group (Figure 3D).

**Figure 3.**
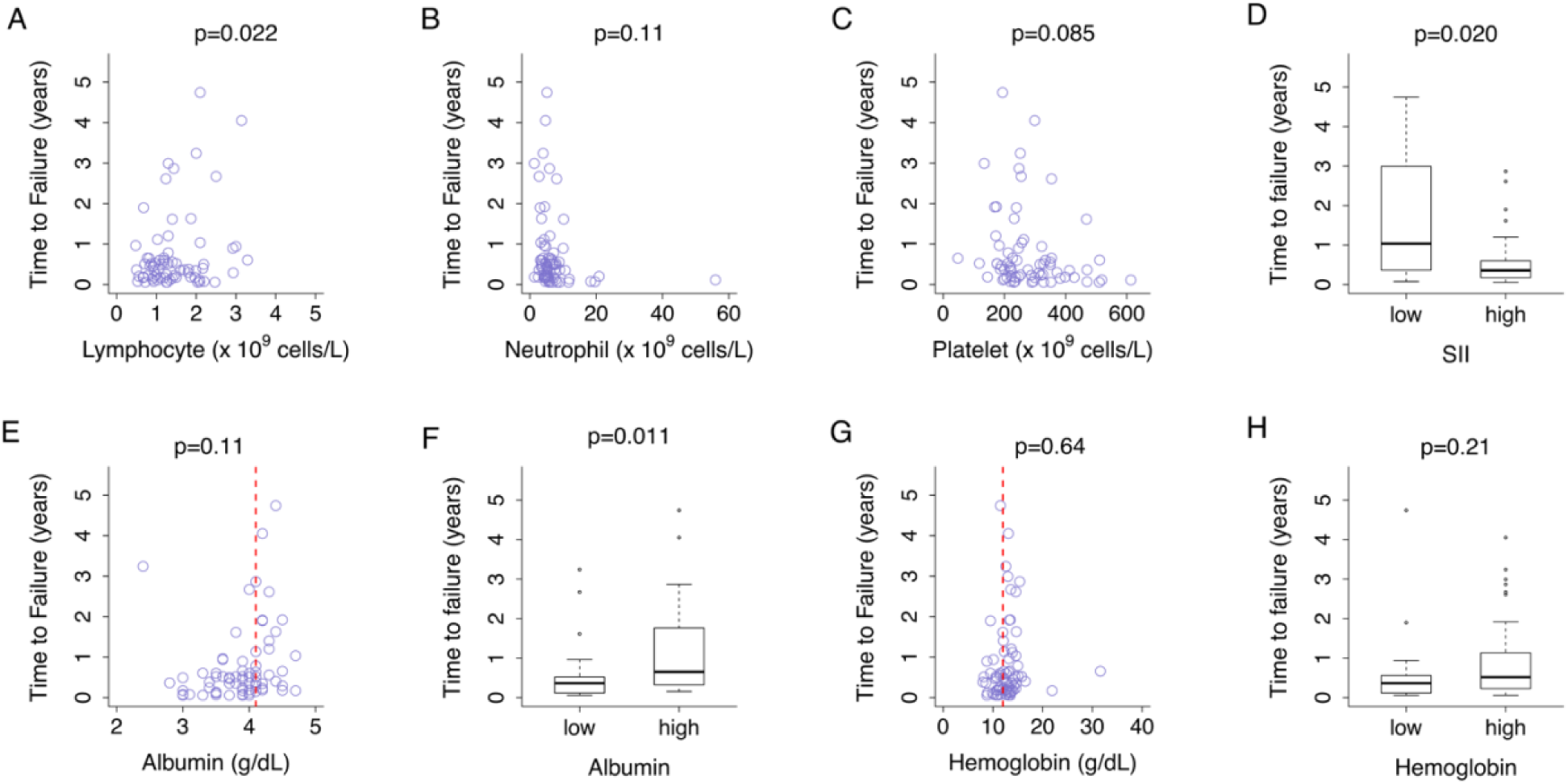
Treatment response and blood biomarkers. **A.** Among 71 patients, a higher lymphocyte count at the start of immunotherapy correlated with longer time to treatment failure (Pearson correlation efficient, *ρ*=0.27 and p=0.022). **B**. Blood neutrophil count at the initiation of immunotherapy negatively correlated with time to failure among 72 patients ( *ρ*=−0.19 and p=0.11). **C**. Platelet count at the initiation of immunotherapy also negatively correlated with time to failure among 71 patients (*ρ*=−0.21 and p=0.085). **D.** Boxplot comparing time to failure for patients with low (<504 ×10^9^ cells/L) and high (≥504 ×10^9^ cells/L) systemic immune-inflammation index (SII). Patients with a low SII had a significantly longer time to failure (mean 1.72 [0.74 – 2.70] years) than those with a high SII (mean 0.52 [0.36 – 0.67] years; t-test, p=0.020). **E.** Association between serum albumin level and time to immunotherapy failure in 68 patients (*ρ*=0.19 and p=0.11). The red dotted line indicates the albumin level of 4.1 g/dL. **F.** Patients with an albumin level of ≥4.1 g/dL (high) had significantly longer time to failure compared to those with an albumin level of <4.1 g/dL (low) (mean 1.19 [0.72 – 1.67] vs. 0.52 [0.31 – 0.72] years; t-test, p=0.011). **G.** Time to immunotherapy failure against hemoglobin level at the onset of immunotherapy among 75 patients (*ρ*=0.054 and p=0.64). The red dotted line denotes the hemoglobin level of 12 g/dL. **H**. Time to failure for patients with low (<12 g/dL) and high (≥12 g/dL) hemoglobin counts. The high hemoglobin group had a longer time to failure than the low hemoglobin group (0.88 [0.61 – 1.15] vs. 0.58 [0.19 – 0.98] years; t-test, p=0.21).

Patients with an albumin level of ≥4.1 g/dL had significantly longer time to failure than those with an albumin level of < 4.1 g/dL, with an AUC of 78% with 80% sensitivity and 72% specificity (Figures 3E and 3F). The high hemoglobin group had a longer time to failure than the low hemoglobin group (AUC 62%; Figures 3G and 3H).

### Immunotherapy, metastasis, and gene mutations

Individuals with bone metastasis prior to initiating immunotherapy had shorter times to treatment failure compared to those without (Figure 4A). We observed a trend towards reduced efficacy in patients with pleura or liver metastasis (Figures 4B and 4C). Collectively, patients with bone, pleural, or liver metastasis had shorter times to failure than those without any of these metastases (Figure 4D). A single indicator of bone, pleura, or liver metastasis marked 71% of AUC. We then compared time to failure in patients with and without non-regional lymph node, brain, contralateral lung, or other location metastasis (Figures 4E-4H).

**Figure 4.**
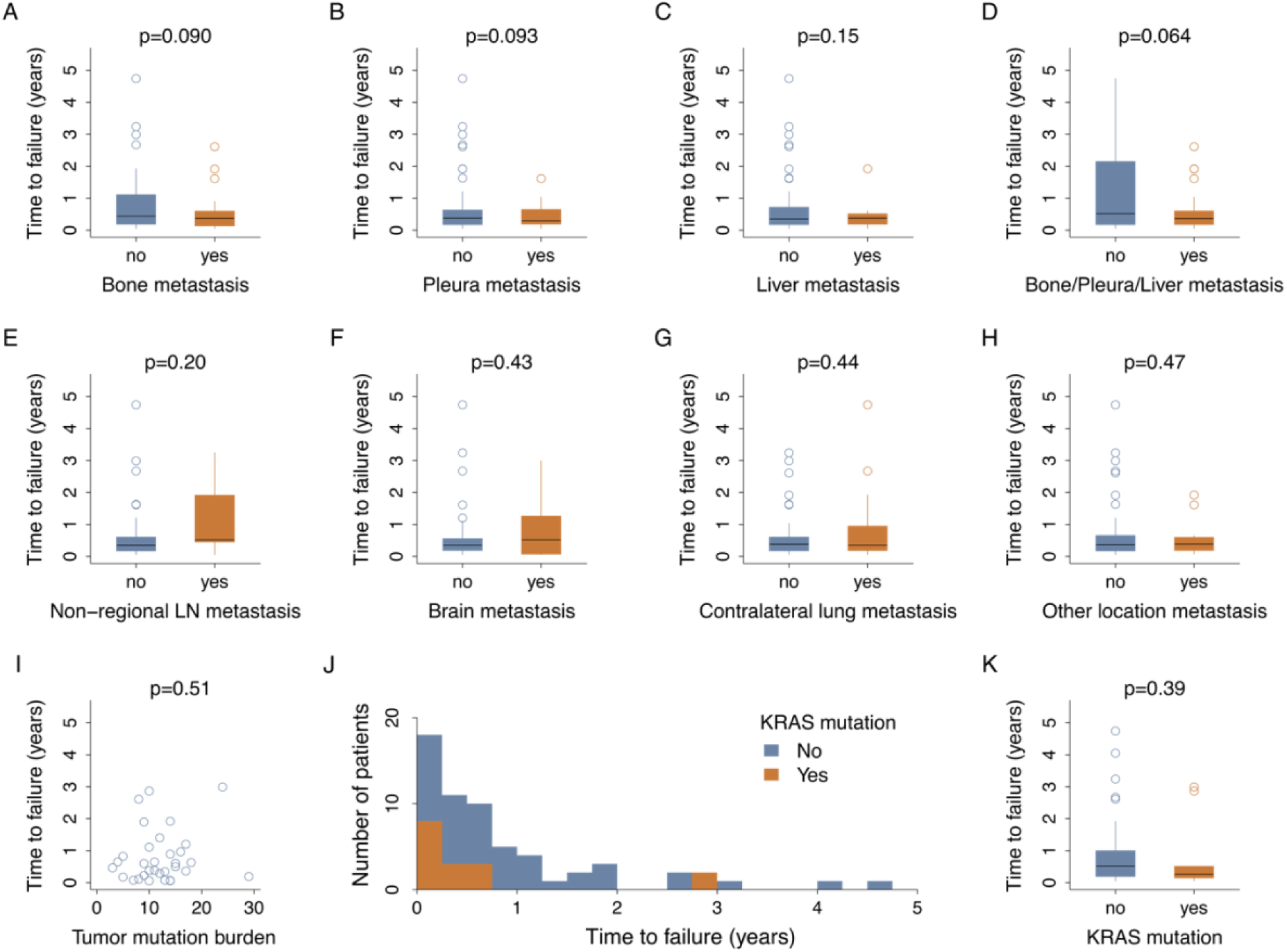
Metastasis, tumor mutation burden, KRAS mutation, and immunotherapy outcome. **A.** Boxplot illustrating the distribution of time to treatment failure for patients without bone metastasis (n=30, 0.90 [0.47 – 1.32] years) and those with bone metastasis (n=34, 0.50 [0.30 – 0.69] years). A shorter time to failure was observed in patients with bone metastasis, and the difference was marginally significant (t-test, p=0.090). **B.** Boxplot comparing time to treatment failure in patients without pleura metastasis (n=47, 0.76 [0.47 – 1.06] years) and those with pleura metastasis (n=17, 0.46 [0.25 – 0.67] years). **C.** Comparison of time to treatment failure between patients without liver metastasis (n=51, 0.74 [0.47 – 1.01] years) and those with liver metastasis (n = 13, 0.47 [0.18 – 0.75] years). **D.** Patients with metastasis in bone, pleura, or liver experienced a shorter time to failure (n=48, 0.50 [0.35 – 0.65] years) than patients without these metastases (n=16, 1.23 [0.47 – 2.00] years). The observed difference was marginally significant (t-test, p=0.064). **E.** Boxplot comparing time to treatment failure in patients without non-regional lymph node metastasis (n=51, 0.60 [0.36 – 0.84] years) and those with non-regional lymph node metastasis (n=13, 1.02 [0.39 – 1.65] years). **F.** Comparison of time to treatment failure between patients without brain metastasis (n=45, 0.62 [0.36 – 0.89] years) and with brain metastasis (n=19, 0.83 [0.38 – 1.27] years). **G.** Patients without contralateral lung metastasis had a mean time to treatment failure of 0.62 [0.39 – 0.85] years (n=45), compared to 0.84 [0.28 – 1.41] years for those with contralateral lung metastasis (n=19). **H.** Comparison of time to treatment failure between patients without metastasis at other locations including kidney, adrenal gland, colon, neck, and thyroid (n=51, 0.72 [0.44 – 0.99] years) and those with metastasis at other locations (n=13, 0.56 [0.22 – 0.91] years). **I.** Scatter plot of time to treatment failure and tumor mutation burden among 32 NSCLC patients. **J.** Distribution of time to immunotherapy failure for NSCLC patients with (n=16, brown) and without (n=59, blue) KRAS mutations. **K.** Boxplot comparison of time to failure between patients with and without KRAS mutations. The average time to failure was higher for individuals without KRAS mutations than for those with KRAS mutations (0.84 [0.58 – 1.09] years vs. 0.61 [0.11 – 1.10] years).

We next investigated the relationship between oncogene mutations and treatment outcomes. Tumor mutation burden was measured in only 32 patients, and no association with treatment efficacy was observed (Figure 4I). Patients with KRAS mutations (n = 16), including G12C and Q61H, experienced immunotherapy failure 84 days sooner on average than those without KRAS mutations (n = 59), though this difference was not statistically significant (Figures 4J and 4K). A lower proportion of patients with KRAS mutations achieved time on treatment exceeding 1 year (12.5%) compared to those without these mutations (25.4%), yielding an AUC of 56%.

### LCatlas prediction accuracy

LCatlas (available at www.lcatlas.org) is a logistic regression based clinical decision support system that predicts first-line immunotherapy outcome and combination therapy (immunotherapy plus chemotherapy) outcome – classifying responses as either extended benefit (time on treatment ≥1 year) or early progression (treatment failure <1 year). The application takes nine clinical and demographic variables as input: gender, smoking history, bone/pleura/liver metastasis, PD-L1 tumor staining percentage, albumin level, hemoglobin level, lymphocyte count, neutrophil count, and platelet count. Lymphocyte, neutrophil, platelet counts are combined into SII, resulting in a logistic regression model processing seven categorical variables. Figure 5A illustrates an example input and output from LCatlas web application.

**Figure 5.**
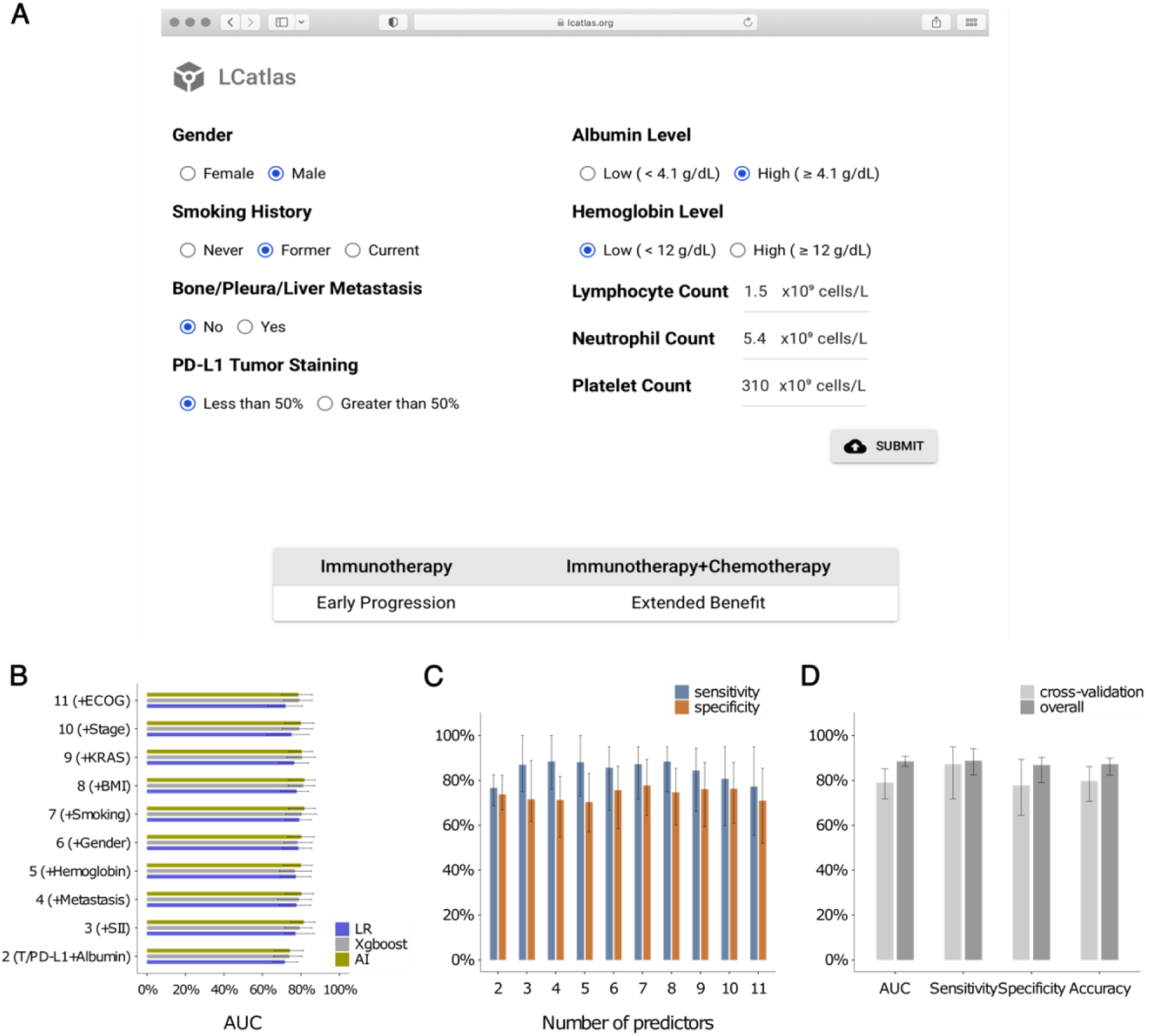
LCaltas: input, output, and evaluation metrics. **A.** LCatlas allows users to input nine variables (gender, smoking history, bone/pleura/liver metastasis, PD-L1 tumor staining percentage, albumin level, hemoglobin level, lymphocyte count, neutrophil count, and platelet count). Upon submission, the model predicts first-line immunotherapy and combination therapy (immunotherapy plus chemotherapy) outcomes – either extended benefit (time on treatment ≥1 year) or early progression (<1 year) – displayed directly on the input page. **B.** Cross-validation AUC (area under the curve) values were assessed as the number predictors increased from 2 to 11 for logistic regression (LR), Xgboost, and TabTransformer AI models in 79 NSCLC patients. The base model included a composite variable of treatment type and PD-L1 tumor staining percentage (T/PD-L1) and albumin level. Subsequent models incorporated the following additional predictors: SII (3 predictors), bone, pleura, or liver metastasis (4 predictors), hemoglobin level (5 predictors), gender (6 predictors), smoking history (7 predictors), BMI (8 predictors), KRAS mutation status (9 predictors), clinical stage (10 predictors), and ECOG score (11 predictors). **C.** Cross-validation sensitivity and specificity were evaluated as the number of predictors increased from 2 to 11. The sum of sensitivity and specificity was maximized using the seven predictors with LCatlas’ nine variables. **D.** LCatlas performance, measured by AUC, sensitivity, specificity, and accuracy with 95% confidence interval, was evaluated in 79 NSCLC patients. Results are presented for overall performance (light grey) and cross-validation (dark grey).

We determined LCatlas predictors by assessing cross-validation accuracy using data from 79 NSCLC patients at the Los Angeles General Medical Center and the University of Southern California Norris Comprehensive Cancer Center (Figure 5B and Supplementary Table S2). The seven predictor model yielded a cross-validation AUC of 79% [72% - 85%] with 87% [72% - 95%] sensitivity, 78% [64% - 89%] specificity, and accuracy of 80% [71% - 86%] (Figures 5B-5D). Both a machine learning (xgboost) and an AI model (TabTransformer) demonstrated comparable performance (80% and 82% AUC each) in cross-validation with the seven predictors (Figure 5B, Supplementary Tables S3 and S4). As shown in Figure 5D, LCatlas achieved 89% [86% - 91%] overall AUC with 89% [82% - 94%] sensitivity, 87% [79% - 90%] specificity, and 87% [82% - 90%] accuracy (Supplementary Table S5).

LCatlas analysis of 79 NSCLC patients showed that higher albumin levels (≥4.1 g/dL) were significantly associated with extended benefit of first-line immunotherapy (odds ratio = 16.1 [2.78 – 93.6]; Table 1 and Supplementary Table S6). Higher hemoglobin levels (≥12 g/dL) and a history of smoking were also associated with extended benefit (odds ratio = 3.07 [0.29 – 33.0] and 2.93 [0.24 – 36.3], respectively). In contrast, a SII greater than 504 ×10^9^ cells/L was associated with early progression of first-line immunotherapy (odds ratio = 0.25 [0.038 – 1.62]). The presence of bone/pleura/liver metastasis and female sex were also associated with early progression, with odds ratios of 0.27 [0.048 – 1.55] and 0.16 [0.026 – 1.02], respectively.

LCatlas’ predictive ability was demonstrated in a patient who achieved over two and a half years of combination therapy duration with PD-L1 staining <50%. This patient had both positive (male, positive smoking history, no bone/pleura/liver metastasis, high hemoglobin) and negative (low albumin level and low SII) impact predictors. LCatlas correctly predicted extended benefit with combination therapy but predicted early progression with immunotherapy alone due to the altered composite predictor of treatment type and PD-L1 tumor staining percentage. Conversely, another patient with both positive (male, positive smoking history, high SII) and negative (bone/pleura/liver metastasis, low albumin level, low hemoglobin level) impact predictors was predicted to have early progression with both immunotherapy alone and combination therapy. LCatlas prediction was consistent with this patient’s outcome (133 days to failure). Collectively, our findings suggest that effective decision-making regarding immunotherapy versus combination therapy may necessitate an integrative approach incorporating diverse clinical and demographic variables.

We further validated LCatlas using a separate validation cohort of 28 NSCLC patients. The model correctly predicted treatment outcomes for 23 patients, yielding an overall accuracy of 82%. Among the two patients who received immunotherapy for more than one year, LCatlas predicted extended benefit for one and early progression for the other. Additionally, of the 26 patients whose treatment failed within a year, 22 were correctly identified as early progressions.

## DISCUSSION

We developed LCatlas, a user-friendly point-of-care tool that predicts immunotherapy response in NSCLC patients. A key advantage of LCatlas over existing models is its ease of accessibility to clinicians and easily implementable with standard vitals and lab tests, supporting individualized therapy decisions in clinical practice. Using nine routinely collected demographic, clinical, and molecular variables, LCatlas classifies patients into two groups: those likely to benefit from treatment for more than 1 year and those at higher risk of treatment failure within 1 year. In a real-world cohort of 79 NSCLC patients, LCatlas achieved an AUC of 89% (89% sensitivity and 87% specificity), highlighting its potential as a valuable clinical resource. The underlying logistic regression model showed an 79% AUC through our cross-validation tests, which was comparable to alternatives including machine learning (xgboost) and AI (TabTransformer) models. In a separate validation cohort (n=28), the model’s overall prediction accuracy was 82%.

Compared to a simpler three-variable model [19], LCatlas achieved higher sensitivity (87% vs. 58%) without compromising specificity (78%) in cross-validation tests. Furthermore, LCatlas demonstrated greater accuracy than a deep learning approach utilizing whole slide hematoxylin and eosin–stained images and PD-L1 score [16]. While a direct comparison was not feasible, LCatlas also outperformed other deep learning-based models using CT-based radiomics [17] and a machine-learning model using cytokine profiles [18] which showed C-indices of 75% and 76%, respectively.

We separately predicted outcomes for immunotherapy and combination therapy by combining first-line therapy type with baseline PD-L1 tumor staining level. We did not observe any associations between PD-L1 level and treatment duration suggesting the limitations of relying heavily on PD-L1 to predict immunotherapy treatment response [27, 28]. Additionally, first-line therapy type – whether immunotherapy alone or in combination with chemotherapy – did not significantly impact treatment failure time. However, combining these two parameters [8] revealed a mean difference of greater than three months in time to treatment failure between groups. LCatlas utilizes this composite variable to independently predict responses to immunotherapy and combination therapy based on PD-L1 level. Given the current imbalance in group sizes for this variable (68.4% vs. 19.0%); consequently, expanding the sample size in future studies would further strengthen our approach.

Previous studies have indicated that smoking status may influence immunotherapy effectiveness in NSCLC [29, 30]. Our analysis confirmed this association, demonstrating that patients with a history of smoking – including both former and current smokers – had significantly longer time on treatment compared to those with no smoking history. Consequently, LCatlas incorporates smoking history as a key input variable (categorized as never vs. former/current). It has been proposed that the improved response to immunotherapy in smokers as compared to never-smokers is due to a higher tumor mutational burden and immunogenicity [30]. In addition, we also take gender as an input, as male patients demonstrated better responses to immunotherapy than female patients, consistent with previous reports [31, 32]. The relationship between body mass index (BMI) and immunotherapy efficacy in NSCLC is unclear [33], with conflicting reports of improved outcomes in patients with higher BMI [34, 35] or no association [36, 37]. In our cohort, individuals with BMI ≥25 had a higher rate of time on treatment lasting longer than 1 year compared to those with BMI <25. However, BMI was not included as a predictor in LCatlas, as its addition did not improve cross-validation accuracy.

Our tool included variables correlated with time to treatment failure in our cohort. The neutrophil-to-lymphocyte ratio (NLR) has been extensively investigated as a prognostic indicator for assessing the likelihood of successful treatment with immune checkpoint inhibitors (ICIs) across various cancer types [38, 39]. More recently, lower systemic inflammation-immune index (SII) values have been linked to better immunotherapy responses in patients with advanced NSCLC [25]. These observations were also confirmed in our cohort, and LCatlas uses SII – a single predictor derived from lymphocyte, neutrophil, and platelet counts. Albumin level was another molecular variable we examined, demonstrating a strong association with immunotherapy outcome in our cohort, consistent with prior studies [40, 41]. A positive correlation coefficient was observed between hemoglobin counts and time to treatment failure, with 62% AUC, as previously reported [42]. We therefore included categorical inputs for both variables.

LCatlas considers patients’ metastasis status; for those with liver, bone, or pleural metastasis, a trend toward reduced efficacy was observed (AUC=71%). Our observation aligns with previous studies reporting the adverse impact of liver metastasis on immunotherapy [44, 45] and reduced immunotherapy effectiveness with bone metastasis [46, 47]. Organ-specific resistance to immunotherapy has been previously reported, potentially due to the immunosuppressive microenvironment observed in liver and bone metastasis [48]. Associations between pleural metastasis and lower treatment response have been also documented [49]. In this study, we combined bone, liver, and pleural metastasis into a single predictor indicating metastasis in any of these organs, which showed an odds ratio of 0.27 [0.05 − 1.55] for predicting immunotherapy outcome.

Patients with KRAS mutations tended to experience treatment failure sooner (84 days on average) than those without, although this association was not statistically significant. Our analysis suggests a potential link between KRAS mutations and reduced treatment efficacy, however, this study is limited by the absence of data on STK11, KEAP1, and NFE2L2 gene mutations along with KRAS G12D mutations and prevalence of KRAS G12C mutations, factors that may modulate the role of immunotherapy in patients with KRAS alteration [50, 51]. Given that KRAS mutation information did not improve the model’s predictive power, it was not included. Additionally, we did not observe an association between tumor mutation burden (measured from only 32 patients) and response to immunotherapy. Associations between tumor mutation burden and immunotherapy response have been reported [52, 53]; however, there has been a lack of consensus regarding the predictive value of specific tumor mutation burden levels for response to immunotherapy [54].

This study is limited by the relatively small number of patients from two study sites, which may restrict the generalizability of our findings. Confirmation in an independent population is needed, and the public availability of the tool online will facilitate this effort. Another limitation is that our feature selection process included limited genomic information and did not include other potential biomarkers such as co-mutations, cytokine profiles [18], circulating tumor DNA (ctDNA) on diagnosis [55], and immune metabolic biomarkers [56].

However, this limitation can also be viewed as a strength. LCatlas leverages routinely collected input variables to identify patients likely to benefit from immunotherapy or immunotherapy plus chemotherapy. This ready accessibility simplifies patient communication regarding treatment rationale and supports evidence-based decisions. Unlike tests requiring additional procedures or incurring significant out-of-pocket costs – many of which are not FDA approved – LCatlas avoids these burdens. Given existing delays in standard-of-care workup – including next-generation sequencing (NGS), PD-L1 testing, and completing disease staging – LCatlas may allow clinicians to inform treatment decisions immediately, avoiding further delays associated with additional testing.

## Author Contributions

S.Y.P., R.H., J. N., and H.L. conceived and designed the study. A.L. and Y.W. compiled the medical records. S.Y.P. and H.L. developed the algorithm and web application. S.Y.P. and H.L. performed data analysis. All authors contributed to the interpretation of results and provided feedback on manuscript drafts. All authors read and approved the final manuscript. All authors were responsible for the decision to submit for publication.

## Institutional Review Board Statement

Access to patient records was approved by the University of Southern California Institutional Review Board (IRB HS-15-00200).

## Informed Consent Statement

Given the retrospective nature of the study, informed consent was waived.

## Data availability statement

LCatlas is publicly available at https://www.lcatlas.org.

## REFERENCES

1. Luo G, Zhang Y, Rumgay H, Morgan E, Langselius O, Vignat J, et al. Estimated worldwide variation and trends in incidence of lung cancer by histological subtype in 2022 and over time: a population-based study. Lancet Respir Med. 2025;13(4):348–63.

2. Ganti AK, Klein AB, Cotarla I, Seal B, Chou E. Update of Incidence, Prevalence, Survival, and Initial Treatment in Patients With Non-Small Cell Lung Cancer in the US. JAMA Oncol. 2021;7(12):1824–32.

3. Velcheti V, Rai P, Kao YH, Chirovsky D, Nunes AT, Liu SV. 5-Year Real-World Outcomes With Frontline Pembrolizumab Monotherapy in PD-L1 Expression >/= 50% Advanced NSCLC. Clin Lung Cancer. 2024;25(6):502–8 e3.

4. Hui R, Garon EB, Goldman JW, Leighl NB, Hellmann MD, Patnaik A, et al. Pembrolizumab as first-line therapy for patients with PD-L1-positive advanced non-small cell lung cancer: a phase 1 trial. Annals of oncology : official journal of the European Society for Medical Oncology / ESMO. 2017;28(4):874–81.

5. Gettinger S, Rizvi NA, Chow LQ, Borghaei H, Brahmer J, Ready N, et al. Nivolumab Monotherapy for First-Line Treatment of Advanced Non-Small-Cell Lung Cancer. J Clin Oncol. 2016;34(25):2980–7.

6. Nikoo M, Rabiee F, Mohebbi H, Eghbalifard N, Rajabi H, Yazdani Y, et al. Nivolumab plus ipilimumab combination therapy in cancer: Current evidence to date. Int Immunopharmacol. 2023;117:109881.

7. Zhang L, Sun L, Zhou Y, Yu J, Lin Y, Wasan HS, et al. Association of Survival and Immune-Related Adverse Events With Anti-PD-1/PD-L1 and Anti-CTLA-4 Inhibitors, Alone or Their Combination for the Treatment of Cancer: A Systematic Review and Meta-Analysis of 13 Clinical Trials. Front Oncol. 2021;11:575457.

8. Reck M, Rodriguez-Abreu D, Robinson AG, Hui R, Csoszi T, Fulop A, et al. Pembrolizumab versus Chemotherapy for PD-L1-Positive Non-Small-Cell Lung Cancer. N Engl J Med. 2016;375(19):1823–33.

9. Sutton RT, Pincock D, Baumgart DC, Sadowski DC, Fedorak RN, Kroeker KI. An overview of clinical decision support systems: benefits, risks, and strategies for success. NPJ Digit Med. 2020;3:17.

10. Duchemann B, Remon J, Naigeon M, Cassard L, Jouniaux JM, Boselli L, et al. Current and future biomarkers for outcomes with immunotherapy in non-small cell lung cancer. Transl Lung Cancer Res. 2021;10(6):2937–54.

11. Zhang J, Song Z, Zhang Y, Zhang C, Xue Q, Zhang G, et al. Recent advances in biomarkers for predicting the efficacy of immunotherapy in non-small cell lung cancer. Front Immunol. 2025;16:1554871.

12. Ricciuti B, Jones G, Severgnini M, Alessi JV, Recondo G, Lawrence M, et al. Early plasma circulating tumor DNA (ctDNA) changes predict response to first-line pembrolizumab-based therapy in non-small cell lung cancer (NSCLC). J Immunother Cancer. 2021;9(3).

13. Wang C, Yu Q, Song T, Wang Z, Song L, Yang Y, et al. The heterogeneous immune landscape between lung adenocarcinoma and squamous carcinoma revealed by single-cell RNA sequencing. Signal Transduct Target Ther. 2022;7(1):289.

14. Oh MS, Abascal J, Rennels AK, Salehi-Rad R, Dubinett SM, Liu B. Tumor Heterogeneity and the Immune Response in Non-Small Cell Lung Cancer: Emerging Insights and Implications for Immunotherapy. Cancers (Basel). 2025;17(6).

15. Saad MB, Al-Tashi Q, Hong L, Verma V, Li W, Boiarsky D, et al. Machine-learning driven strategies for adapting immunotherapy in metastatic NSCLC. Nature communications. 2025;16(1):6828.

16. Rakaee M, Tafavvoghi M, Ricciuti B, Alessi JV, Cortellini A, Citarella F, et al. Deep Learning Model for Predicting Immunotherapy Response in Advanced Non-Small Cell Lung Cancer. JAMA Oncol. 2025;11(2):109–18.

17. Le VH, Minh TNT, Kha QH, Le NQK. Deep Learning Radiomics for Survival Prediction in Non-Small-Cell Lung Cancer Patients from CT Images. J Med Syst. 2025;49(1):22.

18. Wei F, Azuma K, Nakahara Y, Saito H, Matsuo N, Tagami T, et al. Machine learning for prediction of immunotherapeutic outcome in non-small-cell lung cancer based on circulating cytokine signatures. J Immunother Cancer. 2023;11(7).

19. Benzekry S, Grangeon M, Karlsen M, Alexa M, Bicalho-Frazeto I, Chaleat S, et al. Machine Learning for Prediction of Immunotherapy Efficacy in Non-Small Cell Lung Cancer from Simple Clinical and Biological Data. Cancers (Basel). 2021;13(24).

20. Aisner DL, Axtell, A. L., Bauman, J. R., Chang, J. Y. et al. NCCN Guidelines Version 3.2026 Non-Small Cell Lung Cancer. 2025. Available from: https://www.nccn.org/.

21. Reuss JE, Bazhenova L, Ismaila N, Abu Rous F, Ahluwalia A, Alluri K, et al. Therapy for Stage IV Non-Small Cell Lung Cancer Without Driver Alterations: ASCO Living Guideline, 2026.3.0. J Clin Oncol. 2026;44(7):e56–e88.

22. Poei D, Ali S, Thomas JS, Nieva JJ, Hsu RC. Real-World Incidence of Anaplastic Lymphoma Kinase Alterations in Hispanics with Non-Small Cell Lung Cancer at a Large Academic Institution in Los Angeles. Cancer Res Commun. 2025;5(2):277–86.

23. Ettinger DS, Wood DE, Aisner DL, Akerley W, Bauman JR, Bharat A, et al. Non-Small Cell Lung Cancer, Version 3.2022, NCCN Clinical Practice Guidelines in Oncology. J Natl Compr Canc Netw. 2022;20(5):497–530.

24. Nost TH, Alcala K, Urbarova I, Byrne KS, Guida F, Sandanger TM, et al. Systemic inflammation markers and cancer incidence in the UK Biobank. European journal of epidemiology. 2021;36(8):841–8.

25. Feng FS, M. Yao Y., Gao C., Jing Z., Sun C. Predictive Effect of Systemic Immune-Inflammation Index on Immunotherapy in Patients With Advanced Non-Small Cell Lung Cancer. Cancer Control. 2025;32:1–11.

26. Chen TaG, C. XGBoost: A Scalable Tree Boosting System. Proceedings of the 22nd ACM SIGKDD International Conference on Knowledge Discovery and Data Mining. 2016:785--94.

27. Aguilar EJ, Ricciuti B, Gainor JF, Kehl KL, Kravets S, Dahlberg S, et al. Outcomes to first-line pembrolizumab in patients with non-small-cell lung cancer and very high PD-L1 expression. Annals of oncology : official journal of the European Society for Medical Oncology / ESMO. 2019;30(10):1653–9.

28. Incorvaia L, Fanale D, Badalamenti G, Barraco N, Bono M, Corsini LR, et al. Programmed Death Ligand 1 (PD-L1) as a Predictive Biomarker for Pembrolizumab Therapy in Patients with Advanced Non-Small-Cell Lung Cancer (NSCLC). Adv Ther. 2019;36(10):2600–17.

29. Popat S, Liu SV, Scheuer N, Gupta A, Hsu GG, Ramagopalan SV, et al. Association Between Smoking History and Overall Survival in Patients Receiving Pembrolizumab for First-Line Treatment of Advanced Non-Small Cell Lung Cancer. JAMA Netw Open. 2022;5(5):e2214046.

30. Zhao W, Jiang W, Wang H, He J, Su C, Yu Q. Impact of Smoking History on Response to Immunotherapy in Non-Small-Cell Lung Cancer: A Systematic Review and Meta-Analysis. Front Oncol. 2021;11:703143.

31. Wang C, Qiao W, Jiang Y, Zhu M, Shao J, Ren P, et al. Effect of sex on the efficacy of patients receiving immune checkpoint inhibitors in advanced non-small cell lung cancer. Cancer Med. 2019;8(8):4023–31.

32. Conforti F, Pala L, Pagan E, Corti C, Bagnardi V, Queirolo P, et al. Sex-based differences in response to anti-PD-1 or PD-L1 treatment in patients with non-small-cell lung cancer expressing high PD-L1 levels. A systematic review and meta-analysis of randomized clinical trials. ESMO Open. 2021;6(5):100251.

33. Wang Z, Aguilar EG, Luna JI, Dunai C, Khuat LT, Le CT, et al. Paradoxical effects of obesity on T cell function during tumor progression and PD-1 checkpoint blockade. Nat Med. 2019;25(1):141–51.

34. Zhang T, Li S, Chang J, Qin Y, Li C. Impact of BMI on the survival outcomes of non-small cell lung cancer patients treated with immune checkpoint inhibitors: a meta-analysis. BMC cancer. 2023;23(1):1023.

35. Kichenadasse G, Miners JO, Mangoni AA, Rowland A, Hopkins AM, Sorich MJ. Association Between Body Mass Index and Overall Survival With Immune Checkpoint Inhibitor Therapy for Advanced Non-Small Cell Lung Cancer. JAMA Oncol. 2020;6(4):512–8.

36. Cortellini A, Ricciuti B, Vaz VR, Soldato D, Alessi JV, Dall’Olio FG, et al. Prognostic effect of body mass index in patients with advanced NSCLC treated with chemoimmunotherapy combinations. J Immunother Cancer. 2022;10(2).

37. Nie W, Lu J, Qian J, Wang SY, Cheng L, Zheng L, et al. Obesity and survival in advanced non-small cell lung cancer patients treated with chemotherapy, immunotherapy, or chemoimmunotherapy: a multicenter cohort study. Bmc Med. 2024;22(1):463.

38. Sacdalan DB, Lucero JA, Sacdalan DL. Prognostic utility of baseline neutrophil-to-lymphocyte ratio in patients receiving immune checkpoint inhibitors: a review and meta-analysis. Onco Targets Ther. 2018;11:955–65.

39. Alessi JV, Ricciuti B, Alden SL, Bertram AA, Lin JJ, Sakhi M, et al. Low peripheral blood derived neutrophil-to-lymphocyte ratio (dNLR) is associated with increased tumor T-cell infiltration and favorable outcomes to first-line pembrolizumab in non-small cell lung cancer. J Immunother Cancer. 2021;9(11).

40. Chen Y, Liu T, Feng H, Liu T, Zhang J, Wang J, et al. The prognostic role of albumin levels in lung cancer patients receiving third-line or advanced immunotherapy: a retrospective study. Transl Lung Cancer Res. 2024;13(6):1307–17.

41. Guo Y, Wei L, Patel SH, Lopez G, Grogan M, Li M, et al. Serum Albumin: Early Prognostic Marker of Benefit for Immune Checkpoint Inhibitor Monotherapy But Not Chemoimmunotherapy. Clin Lung Cancer. 2022;23(4):345–55.

42. Zhang Z, Zhang F, Yuan F, Li Y, Ma J, Ou Q, et al. Pretreatment hemoglobin level as a predictor to evaluate the efficacy of immune checkpoint inhibitors in patients with advanced non-small cell lung cancer. Ther Adv Med Oncol. 2020;12:1758835920970049.

43. Lee YJ, Park YS, Lee HW, Park TY, Lee JK, Heo EY. Peripheral lymphocyte count as a surrogate marker of immune checkpoint inhibitor therapy outcomes in patients with non-small-cell lung cancer. Sci Rep. 2022;12(1):626.

44. Qin BD, Jiao XD, Liu J, Liu K, He X, Wu Y, et al. The effect of liver metastasis on efficacy of immunotherapy plus chemotherapy in advanced lung cancer. Crit Rev Oncol Hematol. 2020;147:102893.

45. Qu FJ, Zhou Y, Wu S. Progress of immune checkpoint inhibitors therapy for non-small cell lung cancer with liver metastases. British journal of cancer. 2024;130(2):165–75.

46. Landi L, D’Inca F, Gelibter A, Chiari R, Grossi F, Delmonte A, et al. Bone metastases and immunotherapy in patients with advanced non-small-cell lung cancer. J Immunother Cancer. 2019;7(1):316.

47. Zhu YJ, Chang XS, Zhou R, Chen YD, Ma HC, Xiao ZZ, et al. Bone metastasis attenuates efficacy of immune checkpoint inhibitors and displays "cold" immune characteristics in Non-small cell lung cancer. Lung Cancer. 2022;166:189–96.

48. Abbott AG, Meyers DE, Elmi-Assadzadeh G, Stukalin I, Marro A, Puloski SKT, et al. Effectiveness of immune checkpoint inhibitor therapy on bone metastases in non-small-cell lung cancer. Front Immunol. 2024;15:1379056.

49. Epaillard N, Benitez JC, Gorria T, Fabre E, Riudavets M, Reyes R, et al. Pleural effusion is a negative prognostic factor for immunotherapy in patients with non-small cell lung cancer (NSCLC): The pluie study. Lung Cancer. 2021;155:114–9.

50. Knetki-Wroblewska M, Wojas-Krawczyk K, Krawczyk P, Krzakowski M. Emerging insights into STK11, KEAP1 and KRAS mutations: implications for immunotherapy in patients with advanced non-small cell lung cancer. Transl Lung Cancer Res. 2024;13(12):3718–30.

51. Tang D, Kang R. NFE2L2 and ferroptosis resistance in cancer therapy. Cancer Drug Resist. 2024;7:41.

52. Hellmann MD, Ciuleanu TE, Pluzanski A, Lee JS, Otterson GA, Audigier-Valette C, et al. Nivolumab plus Ipilimumab in Lung Cancer with a High Tumor Mutational Burden. N Engl J Med. 2018;378(22):2093–104.

53. Ricciuti B, Wang X, Alessi JV, Rizvi H, Mahadevan NR, Li YY, et al. Association of High Tumor Mutation Burden in Non-Small Cell Lung Cancers With Increased Immune Infiltration and Improved Clinical Outcomes of PD-L1 Blockade Across PD-L1 Expression Levels. JAMA Oncol. 2022;8(8):1160–8.

54. Strickler JH, Hanks BA, Khasraw M. Tumor Mutational Burden as a Predictor of Immunotherapy Response: Is More Always Better? Clinical cancer research : an official journal of the American Association for Cancer Research. 2021;27(5):1236–41.

55. Bartolomucci A, Nobrega M, Ferrier T, Dickinson K, Kaorey N, Nadeau A, et al. Circulating tumor DNA to monitor treatment response in solid tumors and advance precision oncology. NPJ Precis Oncol. 2025;9(1):84.

56. Hatae R, Chamoto K, Kim YH, Sonomura K, Taneishi K, Kawaguchi S, et al. Combination of host immune metabolic biomarkers for the PD-1 blockade cancer immunotherapy. JCI Insight. 2020;5(2).

57. Forget P, Khalifa C, Defour JP, Latinne D, Van Pel MC, De Kock M. What is the normal value of the neutrophil-to-lymphocyte ratio? BMC Res Notes. 2017;10(1):12.

