## Supporting Information for "LCatlas: A Point-Of-Care Tool for Predicting Immunotherapy Outcomes in Non-Small Cell Lung Cancer"

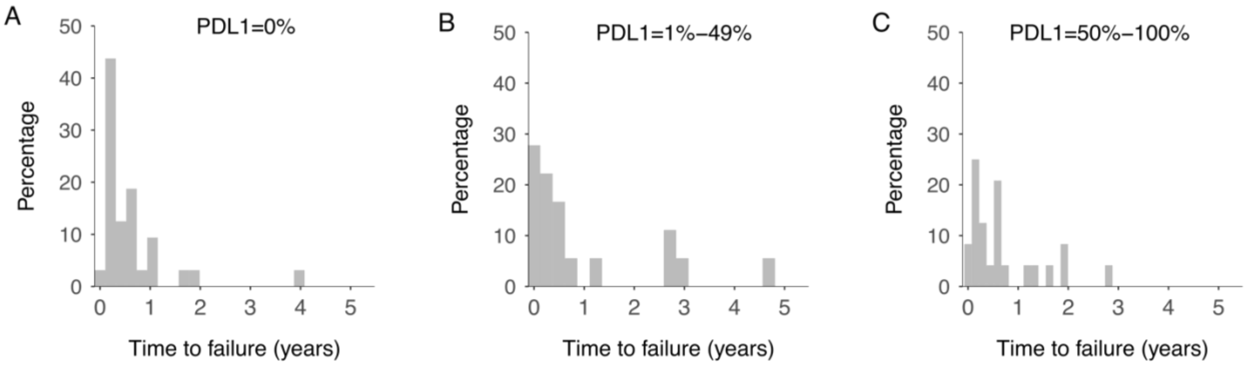


**Supplementary Figure S1. Distribution of time to failure stratified by PD-L1 expression in tumor cells at treatment initiation**. PD-L1 expression group: 0% (n=32, **A**), 1%-49% (n=18, **B**), and 50%-100% (n=24, **C**). Mean time to failure was 0.62 [0.35 – 0.90], 0.98 [0.32 – 1.65], and 0.70 [0.39 – 1.01] years for the 0%, 1-49%, and 50-100% groups, respectively.

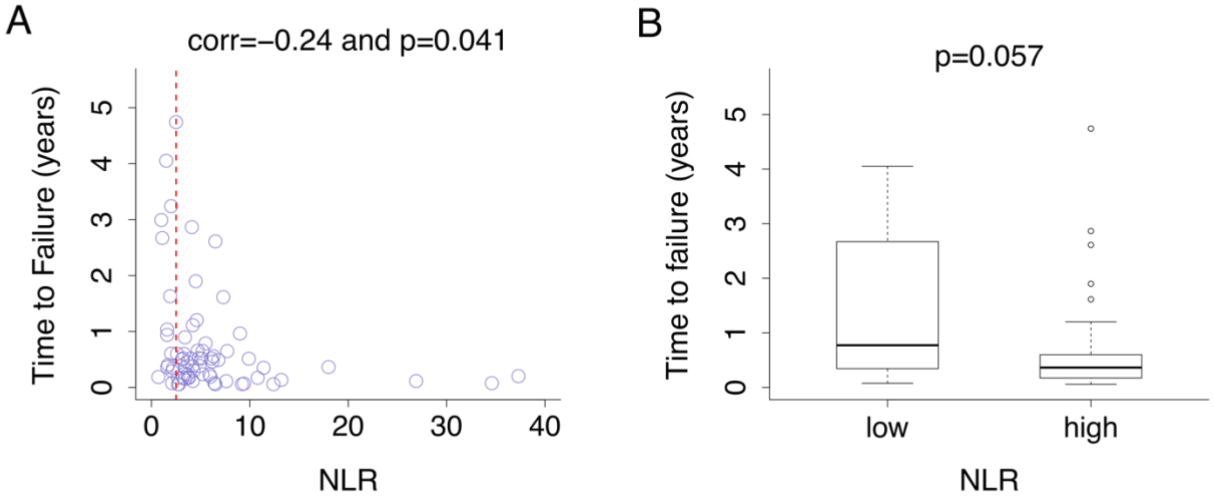


**Supplementary Figure S2. Association between baseline neutrophil-to-lymphocyte ratio (NLR) and time to immunotherapy failure in non-small cell lung cancer patients. A.** Scatter plot of time to immunotherapy failure and NLR at the start of treatment among 71 non-small cell lung cancer patients. The vertical red dotted line denotes an NLR of 2.5. **B**. Boxplot comparing time to failure for patients with low (<2.5) and high (≥2.5) NLR values. The mean time to failure was 1.35 [0.57 – 2.12] years for the low NRL group and 0.58 [0.37 – 0.79] years for the high NLR group. The difference in time to failure between groups was not statistically significant (p=0.057).

| Predictor | Cohort distribution |
|---|---|
| Treatment type/PD-L1 tumor staining | condition1* (53.6%), condition2* (7.1%), NA (39.3%) |
| Albumin | High (39.3%), Low (57.1%), NA (3.6%) |
| SII (Systemic Immune-inflammation Index) | High (39.3%), Low (57.1%), NA (3.6%) |
| Bone/Pleura/Liver metastasis | No (17.9%), Yes (50.0%), NA (32.1%) |
| Hemoglobin | High (64.3%), Low (32.1%), NA (3.6%) |
| Gender | Male (57.1%), Female (42.9), NA (0%) |
| Smoking history | Never (25.0%), Former or current (71.4%), NA (3.6%) |

**Supplementary Table S1. Distribution of variables within the NSCLC validation cohort (n=28).**

*condition1: immunotherapy alone with PD-L1 ≥50% or combination therapy with PD-L1 <50%
*condition2: immunotherapy alone with PDL1 <50% or combination therapy with PD-L1 ≥50%

| Number of Predictors | AUC (Area Under the Curve) | Sensitivity | Specificity | Accuracy | AIC (Akaike Information Criterion) |
|---|---|---|---|---|---|
| 2 (Treatment type/ PD-L1+Albumin) | 72% [66% - 78%] | 77% [69% - 82%] | 74% [67% - 82%] | 74% [69% - 80%] | 57.6 [55.1 - 59.9] |
| 3 (+SII) | 77% [72% - 87%] | 87% [75% - 100%] | 72% [62% - 89%] | 75% [67% - 86%] | 53.8 [51.2 - 56.2] |
| 4 (+Metastasis) | 78% [68% - 85%] | 88% [76% - 100%] | 71% [55% - 82%] | 75% [62% - 84%] | 54.2 [51.3 - 56.5] |
| 5 (+Hemoglobin) | 77% [69% - 85%] | 88% [73% - 100%] | 70% [57% - 83%] | 74% [64% - 83%] | 54.3 [51.5 - 57.5] |
| 6 (+Gender) | 79% [70% - 86%] | 86% [67% - 95%] | 76% [58% - 86%] | 78% [64% - 85%] | 53.1 [49.7 - 55.7] |
| 7 (+Smoking history) | 79% [72% - 85%] | 87% [72% - 95%] | 78% [64% - 89%] | 80% [71% - 86%] | 53.7 [51.4 - 56.3] |
| 8 (+BMI) | 78% [69% - 84%] | 88% [75% - 95%] | 75% [60% - 85%] | 78% [67% - 85%] | 54.3 [50.9 - 56.9] |
| 9 (+KRAS mutation) | 76% [68% - 84%] | 84% [66% - 94%] | 76% [59% - 88%] | 78% [64% - 87%] | 54.9 [49.7 - 58] |
| 10 (+Stage) | 75% [62% - 84%] | 81% [60% - 95%] | 76% [61% - 88%] | 77% [64% - 87%] | 55.1 [49.9 - 58.5] |
| 11 (+ECOG) | 72% [63% - 81%] | 77% [56% - 95%] | 71% [52% - 85%] | 72% [56% - 84%] | 56.6 [51.7 - 60.7] |

**Supplementary Table S2. Average cross-validation metrics with 95% confidence intervals across 100 runs for logistic regression models with varying number of predictors.**

| Number of Predictors | AUC (Area Under the Curve) | Sensitivity | Specificity | Accuracy |
|---|---|---|---|---|
| 2 (Treatment type/ PD-L1+Albumin) | 74% [66% - 81%] | 77% [64% - 89%] | 75% [68% - 84%] | 76% [71% - 81%] |
| 3 (+SII) | 79% [72% - 86%] | 89% [76% - 100%] | 71% [61% - 82%] | 75% [68% - 81%] |
| 4 (+Metastasis) | 79% [68% - 85%] | 90% [75% - 100%] | 72% [59% - 86%] | 76% [66% - 85%] |
| 5 (+Hemoglobin) | 77% [69% - 85%] | 93% [77% - 100%] | 68% [55% - 82%] | 73% [63% - 81%] |
| 6 (+Gender) | 78% [71% - 86%] | 88% [77% - 98%] | 73% [61% - 85%] | 77% [68% - 85%] |
| 7 (+Smoking history) | 80% [72% - 88%] | 88% [73% - 98%] | 78% [66% - 90%] | 80% [71% - 89%] |
| 8 (+BMI) | 81% [73% - 87%] | 90% [79% - 100%] | 77% [63% - 86%] | 80% [70% - 86%] |
| 9 (+KRAS mutation) | 80% [73% - 87%] | 91% [80% - 100%] | 76% [57% - 87%] | 79% [66% - 87%] |
| 10 (+Stage) | 79% [70% - 86%] | 88% [76% - 100%] | 75% [60% - 86%] | 78% [68% - 86%] |
| 11 (+ECOG) | 79% [71% - 86%] | 89% [73% - 100%] | 75% [63% - 86%] | 78% [70% - 86%] |

**Supplementary Table S3. Average cross-validation metrics with 95% confidence intervals across 100 runs for Xgboost models with varying number of predictors.**

| Number of Predictors | AUC (Area Under the Curve) | Sensitivity | Specificity | Accuracy |
|---|---|---|---|---|
| 2 (Treatment type/ PD-L1+Albumin) | 74% [66% - 81%] | 79% [69% - 89%] | 75% [65% - 85%] | 76% [69% - 82%] |
| 3 (+SII) | 81% [74% - 87%] | 93% [81% - 100%] | 71% [62% - 83%] | 75% [69% - 84%] |
| 4 (+Metastasis) | 80% [72% - 86%] | 92% [82% - 100%] | 72% [61% - 82%] | 76% [69% - 84%] |
| 5 (+Hemoglobin) | 80% [71% - 86%] | 90% [76% - 100%] | 74% [63% - 87%] | 77% [70% - 86%] |
| 6 (+Gender) | 80% [73% - 87%] | 88% [76% - 98%] | 77% [66% - 86%] | 80% [72% - 86%] |
| 7 (+Smoking history) | 82% [74% - 87%] | 87% [76% - 95%] | 82% [74% - 89%] | 83% [77% - 87%] |
| 8 (+BMI) | 82% [73% - 87%] | 89% [77% - 95%] | 80% [69% - 90%] | 82% [73% - 89%] |
| 9 (+KRAS mutation) | 80% [74% - 86%] | 88% [76% - 95%] | 80% [69% - 89%] | 82% [74% - 88%] |
| 10 (+Stage) | 80% [72% - 87%] | 86% [73% - 95%] | 80% [69% - 90%] | 82% [74% - 88%] |
| 11 (+ECOG) | 79% [70% - 86%] | 85% [70% - 95%] | 79% [64% - 90%] | 80% [69% - 87%] |

**Supplementary Table S4. Average cross-validation metrics with 95% confidence intervals across 100 runs for TabTransformer AI models with varying number of predictors.**

| Number of Predictors | AUC (Area Under the Curve) | Sensitivity | Specificity | Accuracy | AIC (Akaike Information Criterion) |
|---|---|---|---|---|---|
| 2 (Treatment type/ PD-L1+Albumin) | 75% [71% - 80%] | 76% [71% - 88%] | 77% [58% - 84%] | 77% [66% - 81%] | 74.0 [68.6 - 78.9] |
| 3 (+SII) | 83% [78% - 87%] | 90% [88% - 94%] | 70% [63% - 77%] | 74% [68% - 80%] | 68.1 [61.5 - 74] |
| 4 (+Metastasis) | 85% [81% - 89%] | 91% [77% - 94%] | 74% [61% - 89%] | 77% [68% - 86%] | 68.0 [61.4 - 74.2] |
| 5 (+Hemoglobin) | 87% [83% - 91%] | 89% [71% - 100%] | 75% [61% - 92%] | 78% [68% - 89%] | 68.3 [61.3 - 74.4] |
| 6 (+Gender) | 88% [85% - 91%] | 88% [82% - 94%] | 82% [76% - 89%] | 84% [79% - 87%] | 67.4 [61.1 - 73] |
| 7 (+Smoking history) | 89% [86% - 91%] | 89% [82% - 94%] | 87% [79% - 90%] | 87% [82% - 90%] | 68.0 [62.8 - 73.1] |
| 8 (+BMI) | 89% [87% - 92%] | 92% [82% - 94%] | 85% [77% - 90%] | 87% [81% - 90%] | 69.3 [63.4 - 74.5] |
| 9 (+KRAS mutation) | 90% [87% - 93%] | 93% [82% - 94%] | 85% [77% - 90%] | 87% [81% - 91%] | 69.8 [63.5 - 75.5] |
| 10 (+Stage) | 90% [87% - 93%] | 92% [82% - 94%] | 87% [79% - 94%] | 88% [82% - 92%] | 70.8 [64.6 - 77.1] |
| 11 (+ECOG) | 90% [87% - 93%] | 92% [82% - 94%] | 86% [79% - 94%] | 88% [82% - 92%] | 72.8 [66.2 - 78.7] |

**Supplementary Table S5. Average overall prediction and fitting metrics with 95% confidence intervals across 1,000 imputations for logistic regression models with varying number of predictors.**

| Predictor | Coefficient | Standard error | Odds ratio with 95% CI | p-value |
|---|---|---|---|---|
| Treatment type/PD-L1 tumor staining | -0.94 | 1.12 | 0.39 [0.04 – 3.50] | 0.40 |
| Albumin | 2.78 | 0.90 | 16.11 [2.78 – 93.57] | 0.002 |
| SII (Systemic Immune-inflammation Index) | -1.40 | 0.96 | 0.25 [0.04 – 1.62] | 0.14 |
| Bone/Pleura/Liver metastasis | -1.30 | 0.89 | 0.27 [0.05 – 1.55] | 0.14 |
| Hemoglobin | 1.12 | 1.21 | 3.07 [0.29 – 33.03] | 0.35 |
| Gender | -1.82 | 0.94 | 0.16 [0.03 – 1.02] | 0.052 |
| Smoking history | 1.08 | 1.28 | 2.93 [0.24 – 36.26] | 0.40 |

**Supplementary Table S6. Summary statistics of LCatlas.**
